# Patterns of physical activity among the students of an Indian university and their perceptions about the curricular content concerned with health

**DOI:** 10.1101/2021.06.10.21258728

**Authors:** Arun Kumar Verma, Girish Singh, Kishor Patwardhan

**Affiliations:** Department of Kriya Sharir, Faculty of Ayurveda, Institute of Medical sciences, BHU, Varanasi; Centre of Biostatistics, Institute of Medical sciences, BHU, Varanasi

**Author notes:** Corresponding Author: Kishor Patwardhan.

**Keywords:** Exercise, Physical Activity, Education, Epidemiology

## Abstract

**Background:** University students are at risk of losing their focus on maintaining healthy levels of physical activity because of their engagements with curricular and co-curricular activities. In India, the physical activity levels of adult population have been reported to be declining in the recent years. However, large studies focusing on university students pertaining to their physical activity are not there in Indian context. ‘Do the curricula in higher education promote physical activity?’ is another question that has not been addressed well.

**Objectives:** Our work aims at describing the physical activity levels of the students in a large public-funded central university located in northern India. The study also aims at capturing the student perceptions about the emphasis their curricular activities receive in connection with leading a physically active lifestyle.

**Methods:** This is a cross-sectional descriptive study and uses International Physical Activity Questionnaire (IPAQ-Long form) to record physical activity among 4586 students. Stratified sampling method was used to enroll the students from each stream (faculty). About 15% of all enrolled students from each faculty were included in the study. The study was conducted in between 2016 and 2019. To capture the student perceptions, we have used a 5-item newly developed scale.

**Results:** 2828 (61.7%) male and 1758 (38.3%) female students participated in the study. The mean age of our sample was 22.34 ± 3.12 years. Our results indicate that about 14.5% of all students in the study fall under ‘Inactive’ category. Further, the perception about the curricular content pertaining to physical activity varied widely between the students of different streams.

**Discussion:** Our sample reported a better physical activity pattern in comparison to the reported overall physical activity levels of adult population of India. Our results also suggest that health-related topics are inadequately represented in many of the streams of higher education in the university.

## Introduction

Patterns of physical activity are undergoing significant change in the recent years among individuals of all age groups across the globe.^1-5^ Literature suggests that these changes are mostly influenced by factors such as changing lifestyles, gender differences, economic status, socio-cultural influences, educational levels, occupational factors and other determinants.^6,7^ Many workers in the field have reported a declining trend in physical activity profile among children, young adults and adults across different societies including India.^8-11^ An increased engagement with virtual games, cell phones, television, computers and social media are possibly some of the important contributing factors to this trend among youth. Increased use of vehicular mode of transportation and reduced involvement in outdoor activities also contribute to this outcome.^12-15^ Further, the incidence of health conditions such as overweight, obesity, coronary artery disease, hypertension, diabetes mellitus, depression etc. are known to have increased among young adults and a suboptimal physical activity has been recognized to be an important factor that is associated with these conditions. ^18-23^

In a study conducted by Indian Council of Medical Research (ICMR), physical activity patterns in adults across India were studied. The study reported that, out of 14227 individuals studied, 54.4% were inactive, while 31.9% were active and 13.7% were highly active.^9^ This trend is a matter of concern as the percentage of inactive population appears to be very significant. There are several studies to show that the physical activity levels among youth are going down in comparison to the recommended levels in many countries.^24-29^ Several studies have focused on the physical activity patterns among university students.^23-28^

However, there are no large and methodically performed studies available to show if the overall physical activity levels are lower than the recommended ones among students of Indian Universities. This question becomes important considering the fact that Universities are the places where health awareness is supposed to be inculcated among the youth and these students are at the risk of losing focus on physical activity because of the burden of curricular activities. A few studies have thrown light on the perceptions of student population towards physical education and health-related information in the curriculum.^29-33^ However, such studies are not available in Indian context. This question becomes important keeping the diverse nature of Indian education system and the curricula.

Hence, we planned this study to understand the physical activity trends among the students of Banaras Hindu University (BHU) and also to capture their perceptions regarding the curricular content related to physical activity. BHU is a public central university located in Varanasi, Uttar Pradesh, established in 1916. It is one of the largest residential universities in Asia. BHU is organized into 6 institutes and 14 faculties (streams) and 144 departments. The total student enrolment at the university is around 30,000 and this number represents almost all states of India along with a few foreign countries.

### Objectives of the study

The primary objective of the study was to understand the proportion of students of Banaras Hindu University (BHU) who fall under different categories of physical activity, i.e., physically inactive, active and highly active. The study also aimed at comparing the physical activity profiles of students from different faculties of BHU. Throughout the study we aimed at understanding the differences in the physical activity profiles with respect to age and gender. Another objective of the study was to map views and opinions of the students regarding the information and motivation they receive in their respective faculties and departments as a part of their routine curricular activities to keep themselves physically active.

## Methods

### Study Design and Sampling

This is a cross-sectional survey study wherein a stratified sampling technique was employed. Individual stream (e.g., Humanities, Science, Social Science, Medicine, Ayurveda etc.) was considered as one stratum. We collected the details of the total number of students registered in each of the 16 streams from the offices the respective deans. It was decided to include about 15% of all the students from each stream considering the time and other limitations. This meant approximately 4600 students, which was thought to be sufficient enough to draw meaningful conclusions.

### Tools used in the study

To record the physical activity profiles of the students, we used the International Physical Activity Questionnaire-Long Version (IPAQ-L).^34,35^ This tool has been developed by IPAQ group and is a widely used in large surveys. This tool employs an indirect method of measuring physical activity based on the recall of one’s activities during last one week. The purpose of this tool is to provide a common instrument that can be used to obtain internationally comparable data on health–related physical activity. Further, a newly developed 5-item questionnaire was used to record the opinions and views of the student population. This tool was designed to capture the perceptions of the students regarding the encouragement they receive in their respective faculties and departments to keep themselves physically active.

### Translation and Re-validation of IPAQ-L

The IPAQ-L is available in different languages (English, French, German, Greek etc.) but not in Hindi. Since Hindi is the common language of communication in this part of India, the questionnaire was translated from English to Hindi by an expert. The questionnaire was then back-translated to English and was verified for its accuracy by another team of experts in the department. Suitable corrections were made before the tool was administered. Both the versions of the tool were used in the study to collect data based on student preference.

### Development of a new tool to capture the views and opinions of the student population

A short 5-item questionnaire was developed which was administered to all participants in the study. The statements (items) included in this questionnaire were as follows:

1. The curriculum of my course / courses addresses the topics related to ‘importance of day-to-day physical activity in maintaining health’.
2. My faculty/ department promotes physical activity / sports activities among the students in an organized manner regularly.
3. I consider the sports facilities (playgrounds / sports equipment / sports training) available in my faculty for the students are adequate in general.
4. I keep monitoring my body weight regularly and I am aware of the health consequences of overweight and obesity.
5. I consider that general health-related aspects (such as diet / nutrition / sports etc) are sufficiently addressed in my curriculum.

The options given for each of the questions were in the form of a 5-point Likert Scale: 1=Strongly Agree, 2=Agree, 3= Undecided, 4=Disagree, 5=Strongly Disagree.

### Validation of the new tool

The 5-item scale was first administered to 100 students from the Institute of Agricultural Sciences for the purpose of validation. Cronbach’s Alpha for the scale was 0.725, which falls under the category of acceptable range. Hence, the questionnaire was considered as valid and reliable.

### Data collection and data entry

Ethical clearance was obtained by Institutional Ethics Committee (Reference: No. 2014-15/EC/1323) before starting the study. Investigators collected the data regarding the total number of students registered from different faculties of BHU by writing to the Deans. Since the information contained only numbers and not the list of students, it was decided that the required number of classes be randomly selected and all students of those classes (batches) be administered with the tool. The first author of this paper visited different departments and got permission from concerned heads of the departments to collect the data in leisure hours from different classes. The specific classes were selected by computer generated random sequence method. A written consent was obtained from each of the participant. Though we collected the hard copies of the filled-in questionnaires from the volunteers, to ensure precision and uniformity, we prepared an online form to enter the data. Finally, the data was downloaded in the form of a spreadsheet. The data was collected in between 2016 and 2019. IPAQ-L and the 5-item questionnaire were filled simultaneously by all volunteers.

### Data Analysis

The data analysis to evaluate physical activity patterns was done according to the data processing rules of the IPAQ-L. The major steps involved in this process were data cleaning, excluding the outliers based on the maximum values allowed, ensuring the minimum values for duration of reported activity, truncation of data, calculating MET-minute/week scores for walking, moderate-Intensity and vigorous-Intensity activities and calculating the Total Physical Activity Scores. The final step was to classify the entire sample into categorical data in terms of Low (inactive), Moderate (active) and High (highly active) levels of physical activity.

## Results

The total student strength of BHU was 30667 and upon calculation, 15% of this population is 4600. We collected a sample of 4733. However, after excluding the outliers and erratic entries, the sample that was analyzed included 4586 students. **Table 1** shows the distribution of participants as per their programs of study, gender and age-group. The total number of male and female students included in the study was 2828 (61.7%) and 1758 (38.3%) respectively. Mean age of the sample in the study was 22.34 ± 3.12 years (Males: 22.37 ± 3.13 years and females: 22.29 ± 3.12 years). Out of 4586 students, 3048 (66.4%) were from undergraduate programs, 1406 (30.7%) were from postgraduate programs and 132 (2.9%) were from doctoral level programs.

**Table-1.** Distribution of volunteers as per age-group, gender and program of study. (Percentage expressed is according to rows).

|  |  |  |  |  |  |  |
| --- | --- | --- | --- | --- | --- | --- |
| **Age Group** | **Gender** | **Program of study** |  |  | **Total (based on gender in each age group)** | **Total (in each age group)** |
|  |  | **PhD** | **Postgraduate** | **Undergraduate** |  |  |
| **16-20** | **Female** | - | 7 (1.2%) | 553 (98.8%) | 560 | **1445** |
|  | **Male** | - | 20 (2.3%) | 865 (97.7%) | 885 |  |
| **21-25** | **Female** | 5 (0.5%) | 434 (44.0%) | 547 (55.5%) | 986 | **2539** |
|  | **Male** | 14 (0.9%) | 633 (40.8%) | 906 (58.3%) | 1553 |  |
| **26-30** | **Female** | 17 (9.7%) | 107 (60.8%) | 52 (29.5%) | 176 | **503** |
|  | **Male** | 52 (15.9%) | 163 (49.8%) | 112 (34.3%) | 327 |  |
| **31-35** | **Female** | 11(36.7%) | 16 (53.3%) | 3 (10.0%) | 30 | **82** |
|  | **Male** | 21 (40.4%) | 22 (42.3%) | 9 (17.3%) | 52 |  |
| **36 and above** | **Female** | 4 (66.7%) | 2 (33.3%) | 0 (.0%) | 6 | **17** |
|  | **Male** | 8 (72.7%) | 2 (18.2%) | 1 (9.1%) | 11 |  |
| **Total** | **Female** | **37** | **840** | **1155** | **1758** | **4586** |
|  | **Male** | **95** | **566** | **1893** | **2828** |  |

### Physical Activity Levels

**Table-2** displays the overall distribution of subjects into Low (inactive), Moderate (active) and High (highly active) levels of Physical Activity. In our sample, we noted that about 14.5% of all students fell under Low category of Physical Activity (14.4% among all males and 14.7 among all females), whereas an almost equal proportion, i.e., 14.2% of all students (15.3% of all males and 13.5% of female) fell under Moderate physical activity category. Further, about 71.3% of all students (72.1% among all males and 70% among all females) fell under High level of Physical activity. The difference between physical activity levels for males and females was statistically not significant (χ^2^ =3.237, p = 0.198). Further, the difference was not significant between males and females for any program of study too. **Table-2** also shows the distribution of volunteers into High, Moderate and low levels of physical activity based on their programs of study and gender. Among all PhD scholars 21.2% fall under Low category, 9.1% under Moderate and 69.7% under High category. Among all Postgraduate students, 15.3% fall under Low, 14.2% under Moderate and 70.6% fall under High category. Among Undergraduate students, 13.9% fall under Low category, 14.4% under Moderate and 71.7% fall under High category. The difference between physical activity of students of various programs was not statistically significant (χ^2^ =8.282, p = 0.082).

**Table-2.** Distribution of volunteers into High, Moderate and low levels of physical activity based on programs of study and gender. (Percentage expressed is according to rows)

| Table-2 Distribution of volunteers into High, Moderate and low levels of physical activity based on programs of study and gender. (Percentage expressed is according to rows) |  |  |  |  |  | Comparison between gender and category |
| --- | --- | --- | --- | --- | --- | --- |
| Program of study | Gender | Category |  |  | Total |  |
|  |  | Highly Active | Active | Inactive |  |  |
| Ph. D. | Female | 23 (62.2 %) | 5 (13.5 %) | 9 (24.3 %) | 37 (100 %) | X<sup>2</sup> = 1.760<br>P=0.415 |
|  | Male | 69 (72.6 %) | 7 (7.4 %) | 19 (20.0 %) | 95 (100 %) |  |
|  | Total | 92 (69.7 %) | 12 (9.1 %) | 28 (21.2 %) | 132 (100 %) |  |
| Postgraduate | Female | 380 (67.2 %) | 88 (15.5 %) | 98 (17.3 %) | 566 (100 %) | X<sup>2</sup> = 5.404<br>P=0.067 |
|  | Male | 612(72.9 %) | 111 (13.2 %) | 117 (13.9 %) | 840 (100 %) |  |
|  | Total | 992 (70.5 %) | 199 (14.2 %) | 215 (15.3 %) | 1406 (100 %) |  |
| Undergraduate | Female | 827 (71.6 %) | 176 (15.2 %) | 152 (13.2 %) | 1155 (100 %) | X<sup>2</sup> = 1.522<br>P=0.467 |
|  | Male | 1358 (71.8 %) | 264 (13.9 %) | 271 (14.3 %) | 1893 (100 %) |  |
|  | Total | 2185 (71.7 %) | 440 (14.4 %) | 423 (13.9 %) | 3048 (100 %) |  |
| Total | Male | 2039 (72.1%) | 382 (13.5%) | 407 (14.4%) | 2828 (100 %) | X<sup>2</sup> = 3.237 p = 0.198 |
|  | Female | 1230 (70.0%) | 269(15.3%) | 259 (14.7%) | 1758 (100 %) |  |
| Grand Total |  | 3269 (71.3%) | 651 (14.2%) | 666 (14.5%) | 4586 (100 %) |  |

**Table-3** depicts the distribution of volunteers into High, Moderate and Low categories based on age group. As the table suggests, the number of students in ‘Highly active’ category is highest among lower age groups and the number of students in ‘Inactive’ category is highest among higher age groups. The difference between physical activity of students of different age-groups was statistically significant (χ2 = 35.387, p < 0.000).

**Table-3.** Distribution of volunteers into High, Moderate and Low categories based on age group. Percentage expressed is according to rows.

|  |  |  |  |  |  |  |
| --- | --- | --- | --- | --- | --- | --- |
| Age Group | Gender | Category |  |  | Total | Comparison between males and females |
|  |  | Highly Active | Active | Inactive |  |  |
| 16-20 | Male | 659 (74.5%) | 126 (14.2%) | 100 (11.3%) | 1445<br>(100 %) | $\chi^2=1.327$,<br>p=0.515 |
|  | Female | 402 (71.7%) | 86 (15.4%) | 72 (12.9%) |  |  |
| 21-25 | Male | 1123 (72.3%) | 208 (13.4%) | 222 (14.3%) | 2539<br>(100 %) | $\chi^2=2.988$,<br>p=0.224 |
|  | Female | 686 (69.6%) | 155 (15.7%) | 145 (14.7%) |  |  |
| 26-30 | Male | 218 (66.7%) | 42 (12.8%) | 67 (20.5%) | 503<br>(100 %) | $\chi^2=0.976$,<br>p=0.614 |
|  | Female | 124 (70.5%) | 18 (10.2%) | 34 (19.3%) |  |  |
| 31-35 | Male | 29 (55.8%) | 6 (11.5%) | 17 (32.7 %) | 82<br>(100 %) | $\chi^2=3.663$,<br>p=0.160 |
|  | Female | 16 (53.3%) | 8 (26.7%) | 6 (20%) |  |  |
| 36 and above | Male | 10 (90.9%) | 0 (0%) | 1 (9.1%) | 17<br>(100 %) | $\chi^2=6.783$,<br>**p=0.034** |
|  | Female | 2 (33.4%) | 2 (33.3%) | 2 (33.3%) |  |  |
| Overall comparison of physical activity levels among different age groups: $\chi^2 = 35.387$,<br>p < 0.00 | | | | | | |

### Total Physical Activity MET distribution

**Table-4** depicts the distribution of MET minutes per week under different categories in the form of total walking, total moderate activity, total vigorous activity and total physical activity MET minutes per week. The mean total physical activity MET minutes per week for males was 4678.5 ± 3037.01, and 4321.4 ± 2874.09 for females. Overall mean total physical activity MET minutes per week was 4541.6 ± 2980.35. The difference between the MET min/week among males and females was statistically significant for all categories of physical activity domains reported as suggested by p values.

**Table-4.** Distribution of MET minutes per week under different domains.

| **Table-4. Distribution of MET minutes per week under different domains** |  |  |  |  |
| --- | --- | --- | --- | --- |
| **MET** | **Gender** |  | **Total (N=4586)**<br>**Mean (SD)** | **Mann**<br>**Whitney**<br>**Test**<br>**P value** |
|  | **Female**<br>**(N=1758)**<br>**Mean (SD)** | **Male (N=2828)**<br>**Mean (SD)** |  |  |
| **Total Walking**<br>**MET** | 2107.2 (1334.39) | 2201.2 (1380.27) | 2165.2 (1363.49) | **0.022** |
| **Total Moderate**<br>**MET** | 1387.47<br>(1198.08) | 1498.81<br>(1258.63) | 1456.13<br>(1236.81) | **0.004** |
| **Total Vigorous**<br>**MET** | 826.76 (1665.84) | 978.46 (1760.39) | 920.31 (1726.15) | **0.000** |
| **Total physical**<br>**activity MET** | 4321.4 (2874.09) | 4678.5 (3037.01) | 4541.6 (2980.35) | **0.000** |

### Faculties with Least Active Students

As the **Table-5** suggests, among all the faculties, the Faculty of Ayurveda had a maximum number of least active students, i.e., 41.3%. The following faculties were next in the rank: Education (26.5%), Law (24.6%), Medicine (18.6%), Performing Arts (16.9%), Environmental Science (16.7%), Management (15.9%), Science (14.4%), Arts (13.5%), Social Sciences (12.6%), Agriculture (12.3%), Commerce (12.2%), Women’s College (12.3%), Visual arts (10.9%), Sanskrit Studies (6.7%) and Dental Sciences (2.9%). The difference between physical activity levels in different streams was statistically significant as suggested by p values.

**Table-5.** Distribution of volunteers into High, Moderate and Low categories based on their faculty affiliation.

|  |  |  |  |  |
| --- | --- | --- | --- | --- |
| **Faculty** | **Category** |  |  | **Total** |
|  | **Highly Active** | **Active** | **Inactive** |  |
| Agriculture | 226 (75.1%) | 38 (12.6%) | 37 (12.3%) | 301(100.0%) |
| Arts | 739 (70.9%) | 163 (15.6%) | 140 (13.5%) | 1042 (100.0%) |
| Ayurveda | 33 (41.2%) | 14 (17.5%) | 33 (41.3%) | 80 (100.0%) |
| Commerce | 236 (77.6%) | 31 (10.2%) | 37 (12.2%) | 304 (100.0%) |
| Dental Sciences | 26 (76.5%) | 7 (20.6%) | 1 (2.9%) | 34 (100.0%) |
| Education | 38 (55.9%) | 12 (17.6%) | 18 (26.5%) | 68 (100.0%) |
| Environmental Sciences | 15 (83.3%) | 0 (0.0%) | 3 (16.7%) | 18 (100.0%) |
| Law | 133 (66.8%) | 17 (8.6%) | 49 (24.6%) | 199 (100.0%) |
| Women's College | 333 (75.3%) | 55 (12.4%) | 54 (12.3%) | 442 (100.0%) |
| Management | 42 (66.6%) | 11 (17.5%) | 10 (15.9%) | 63 (100.0%) |
| Medicine | 142 (61.5%) | 46 (19.9%) | 43 (18.6%) | 231 (100.0%) |
| Performing Arts | 127 (67.2%) | 30 (15.9%) | 32 (16.9%) | 189 (100.0%) |
| Sanskrit Studies | 120 (81.1%) | 18 (12.2%) | 10 (6.7%) | 148 (100.0%) |
| Science | 626 (71.1%) | 128 (14.5%) | 127 (14.4%) | 881 (100.0%) |
| Social Sciences | 347 (74.3%) | 61 (13.1%) | 59 (12.6%) | 467 (100.0%) |
| Visual Arts | 86 (72.3%) | 20 (16.8%) | 13 (10.9%) | 119 (100.0%) |
| Total | 3269 (71.3%) | 651 (14.2%) | 666 (14.5%) | 4586 (100.0%) |
| **$\chi^2=126.2$, $p=0.000$** |  |  |  |  |

### Domains of Physical Activity reported

Our sample reported activities for transportation using bicycle (49.18%), walking (91.37%), vigorous housework outside home (26.10%), moderate housework outside home (57.46%), moderate housework inside home (69.97%), vigorous leisure physical activity (40.97%), moderate leisure physical activity (43.46%) and leisure time walking (75.36%). Since all the volunteers were students, they did not report any work-related physical activity.

### Views and Opinions of the Students

**Table-6** shows the responses of the students to each option to the 5-item questionnaire based on gender. The statistically significant difference was observed in the responses for item numbers 1, 3 and 4 among males and females, whereas no statistically significant response was found for the item number 2 and 5.

**Table-6.** shows the number and percentage responses of the students to each option to the 5-item questionnaire based on gender.

|  |  |  |  |  |  |  |  |
| --- | --- | --- | --- | --- | --- | --- | --- |
| Item | Gender | Strongly Agree | Agree | Undecided | Disagree | Strongly Disagree | Chi Square Test |
| 1 | Female (1758) | 451 (25.7%) | 373 (21.2%) | 146 (8.3%) | 503 (28.6%) | 285 (16.2%) | $\chi^2 = 32.68$<br>**p < 0.000** |
|  | Male (2828) | 914 (32.4%) | 626 (22.1%) | 205 (7.2%) | 648 (22.9%) | 435 (15.4%) |  |
| 2 | Female (1758) | 291 (16.5%) | 780 (44.4%) | 204(11.6%) | 314 (17.9%) | 169 (9.6%) | $\chi^2 = 3.55$<br>**p = 0.470** |
|  | Male (2828) | 475 (16.8%) | 1201(42.5%) | 329(11.6%) | 506 (17.9%) | 317 (11.2%) |  |
| 3 | Female (1758) | 392 (22.3%) | 606 (34.5%) | 272 (15.5%) | 326 (18.5%) | 162 (9.2%) | $\chi^2 = 27.36$<br>**p < 0.000** |
|  | Male (2828) | 808 (28.6%) | 879 (31.1%) | 383 (13.5%) | 464 (16.4%) | 294 (10.4%) |  |
| 4 | Female (1758) | 549 (31.1%) | 792 (45.1%) | 128 (7.3%) | 170 (9.7%) | 119 (6.8%) | $\chi^2 = 16.53$<br>**p = 0.002** |
|  | Male (2828) | 997 (35.4%) | 1189(42.0%) | 236 (8.3%) | 270 (9.5%) | 136 (4.8%) |  |
| 5 | Female (1758) | 420 (23.8%) | 501 (28.5%) | 225 (12.8%) | 386 (22.0%) | 226 (12.9%) | $\chi^2 = 6.26$<br>**p = 0.122** |
|  | Male (2828) | 749 (26.5%) | 796 (28.1%) | 319 (11.3%) | 573 (20.3%) | 391 (13.8%) |  |

**Table-7** shows the responses of students to 5-item questionnaire based on the programs in which they are registered. A statistically significant difference between the responses based on the courses registered (UG/PG/PhD) is observed for all 5 items.

**Table-7.** shows the number and percentage responses of students to 5-item questionnaire based on the programs in which they are registered.

|  |  |  |  |  |  |  |  |
| --- | --- | --- | --- | --- | --- | --- | --- |
| Item Number | Program | Strongly Agree | Agree | Undecided | Disagree | Strongly Disagree | Chi Square Test |
| 1 | PhD (132) | 30 (22.7%) | 38 (28.8%) | 13 (9.9%) | 42 (31.8%) | 9 (6.8%) | $\chi^2 = 72.44$<br>**p < 0.000** |
|  | Postgraduate (1406) | 372 (26.5%) | 248 (17.6%) | 98 (7.0%) | 419 (29.8%) | 269 (19.1%) |  |
|  | Undergraduate (3048) | 963 (31.6%) | 713 (23.4%) | 240 (7.9%) | 690 (22.6%) | 442 (14.5%) |  |
| 2 | PhD (132) | 19 (14.4%) | 55 (41.7%) | 23 (17.4%) | 26 (19.7%) | 9 (6.8%) | $\chi^2 = 104.2$<br>**p < 0.000** |
|  | Postgraduate (1406) | 172 (12.3%) | 537 (38.2%) | 172 (12.2%) | 314 (22.3%) | 211 (15.0%) |  |
|  | Undergraduate (3048) | 575 (18.9%) | 1389 (45.6%) | 338 (11.1%) | 480 (15.7%) | 266 (8.7%) |  |
| 3 | PhD (132) | 24 (18.2%) | 51 (38.6%) | 29 (22.0%) | 21 (15.9%) | 7 (5.3%) | $\chi^2 = 54.73$<br>**p < 0.000** |
|  | Postgraduate (1406) | 326 (23.2%) | 422 (30.0%) | 187 (13.3%) | 303 (21.6%) | 168 (11.9%) |  |
|  | Undergraduate (3048) | 850 (27.9%) | 1012 (33.2%) | 439 (14.4%) | 466 (15.3%) | 281 (9.2%) |  |
| 4 | PhD (132) | 52 (39.4%) | 58 (43.9%) | 9 (6.8%) | 10 (7.6%) | 3 (2.3%) | $\chi^2 = 35.60$<br>**p < 0.000** |
|  | Postgraduate (1406) | 425 (30.3%) | 612 (43.5%) | 105 (7.5%) | 151 (10.7%) | 113 (8.0%) |  |
|  | Undergraduate (3048) | 1069 (35.1%) | 1311 (43.0%) | 250 (8.2%) | 279 (9.2%) | 139 (4.5%) |  |
| 5 | PhD (132) | 28 (21.2%) | 38 (28.8%) | 21 (15.9%) | 30 (22.7%) | 15 (11.4%) | $\chi^2 = 44.38$<br>**p < 0.000** |
|  | Postgraduate (1406) | 364 (25.8%) | 336 (23.9%) | 140 (10.0%) | 336 (23.9%) | 230 (16.4%) |  |
|  | Undergraduate (3048) | 777 (25.4%) | 923 (30.3%) | 383 (12.6%) | 593 (19.5%) | 372 (12.2%) |  |

**Table-8** shows the mean scores for each item in each faculty. A mean score lesser than 3 for any item was considered to be indicating a positive perception about the curricular activities leading to an encouraging environment to lead a physically active and healthy lifestyle. A mean score of more than 3 for any item was considered as indicating dissatisfaction towards the curriculum of the faculty with respect to leading a physically active lifestyle. From the table it becomes clear that the faculties of Agriculture Sciences, Arts, Ayurveda, Dental Sciences, Medicine, Performing arts and Science – were the faculties where the mean scores for any of the questions did not exceed 3 or more. Hence, it can be presumed that the students in these faculties get some or the other kind of motivation to lead a physically active lifestyle as a part of their curricular activities.

**Table-8.** Mean scores for each of the five items in the 5-item questionnaire for different faculties.

|  |  |  |  |  |  |  |
| --- | --- | --- | --- | --- | --- | --- |
| **Faculty** | **Number of students** | **Q1**<br>**Mean (SD)** | **Q2**<br>**Mean (SD)** | **Q3**<br>**Mean (SD)** | **Q4**<br>**Mean (SD)** | **Q5**<br>**Mean (SD)** |
| Agriculture | 301 | 2.61 (1.446) | 2.29 (1.222) | 2.40 (1.220) | 1.88 (.981) | 2.55 (1.314) |
| Arts | 1042 | 2.66 (1.569) | 2.71 (1.335) | 2.43 (1.360) | 2.15 (1.177) | 2.71(1.497) |
| Ayurveda | 80 | 2.21 (1.144) | 2.75 (0.921) | 2.51 (1.055) | 2.08 (0.823) | 1.96 (0.878) |
| Commerce | 304 | 3.45 (1.425) | 2.82 (1.316) | 3.07 (1.489) | 1.90 (1.055) | 3.09 (1.417) |
| Dental Sciences | 34 | 2.32 (1.224) | 2.68 (1.121) | 2.68 (1.224) | 1.91 (0.866) | 2.29 (1.142) |
| Education | 68 | 2.93 (1.308) | 2.79 (1.140) | 2.94 (1.170) | 2.09 (1.168) | 3.43 (1.262) |
| Environmental Sciences | 18 | 3.28 (1.364) | 3.22 (1.215) | 3.17 (1.150) | 1.89 (1.132) | 2.72 (1.364) |
| Law | 199 | 3.20 (1.337) | 3.34 (1.203) | 3.31 (1.223) | 2.37 (1.143) | 3.22 (1.238) |
| Women's College | 442 | 3.02 (1.363) | 2.27 (1.096) | 2.30 (1.118) | 2.07 (1.135) | 2.58 (1.276) |
| Management | 63 | 3.56 (1.254) | 2.21 (0.970) | 2.63 (1.140) | 1.95 (0.974) | 2.81 (1.203) |
| Medicine | 231 | 2.19 (1.249) | 2.49 (1.219) | 2.73 (1.361) | 2.03 (1.067) | 2.26 (1.266) |
| Performing Arts | 189 | 1.31 (0.821) | 2.15 (0.686) | 2.26 (1.234) | 2.11 (0.978) | 2.08(1.471) |
| Sanskrit Studies | 148 | 2.58 (1.498) | 2.60 (1.165) | 2.45 (1.039) | 2.30 (1.292) | 2.70 (1.532) |
| Science | 881 | 2.81 (1.439) | 2.48 (1.180) | 2.25 (1.180) | 1.90 (0.964) | 2.51 (1.240) |
| Social Sciences | 467 | 2.93 (1.559) | 3.04 (1.300) | 2.67 (1.394) | 2.57 (1.390) | 3.11 (1.477) |
| Visual Arts | 119 | 2.71 (1.485) | 2.74 (1.374) | 2.96 (1.440) | 2.08 (1.222) | 2.77 (1.362) |
| Total | 4586 | 2.75 (1.492) | 2.62 (1.250) | 2.52 (1.309) | 2.10 (1.136) | 2.69 (1.397) |

## Discussion

### Physical Activity Profiles compared with other studies

This is possibly one of the first studies from India that looks at physical activity levels in a focused way among a large number of university students. According to ICMR study (2014), the total percentage of inactive adults was 54.4% in India.^9^ The percentage of highly active adults was 13.7%. However, the mean age group of this study sample was around 40 years. Since our study sample belongs to a mean age of around 22 years, a true comparison of the results is not possible. However, our results are much encouraging than the ones reported in this study. A study based on the pooled data from 358 population-based surveys from across 168 countries, including 1·9 million participants, reported that the global age-standardized prevalence of insufficient physical activity was 27·5% in 2016, with a difference between sexes of more than 8 percentage points.^2^ In comparison to this, our sample gives a better picture. We report only about 14.5% of inactive student population. Another study conducted among university students in Romania included a total of 333 students, with an age average of 21.05±1.98 years.^27^ According to the results of this study, mean Total Physical Activity MET (TPA-MET) minutes per week are almost comparable with those of our study, especially among female students. The average TPA-MET minutes per week among males was better in their study than in ours. Another study determined the physical activity performed by undergraduate students from 20 to 22 years of age, its frequency and intensity.^28^ The sample consisted of 297 students from the University of Maribor. Their results indicate that 79.8 % students were inactive and hence, our situation in BHU appears to be much better where 71% of students are highly active. In yet another study the investigators investigated the physical activity and quality of life of sports department students and other department students attending university.^29^ A total of 300 university students participated in this study. In comparison with the genders, the total average physical activity score of men was found to be 4938.86 ± 3919.33 MET-minute/week while that of women was found to be 2592.44 ± 2276.82 MET-minute/week. In comparison to these results, female students in our study are much more physically active. According to the results of a study consisting of 200 study subjects, 59% were having a sedentary lifestyle, 27% were moderately active and 14% had vigorously active lifestyle. The study was conducted among the patients attending health training centers in Nagpur and participants’ age ranged from 40 to more than 70 years.^5^ This study reported a significantly increasing trend for sedentary lifestyle with age a finding that is consistent with our results too, though the age range of the subjects in our study was different. A study conducted in urban and rural Vellore city, Tamil Nadu, assessed the prevalence and factors associated with insufficient physical activity among adults aged 30–64 years.^11^ The prevalence of insufficient physical activity was 63.3% in the urban area and 40.6% in the rural area. Though our results cannot be meaningfully compared with this study as the sample characters are different, we report a better physical activity profile.

### Student Perceptions

Our study suggests that the student perceptions vary significantly from one stream of study to another indicating that the curricular activities of all streams do not encourage physically active lifestyle equally. The curricular activities of Agriculture Sciences, Arts, Ayurveda, Dental Sciences, Medicine, Performing arts and Science appear to be encouraging physical activity in one or the other form. This heterogenous perception indicates that there is a need for having a relook at all curricula to see if sufficient emphasis is placed in health-related aspects.

Growing healthcare burden of India is mainly due to increasing prevalence of lifestyle related diseases such as hypertension, obesity, diabetes, depression, metabolic syndrome etc. Increasing use of sugars, fats and other high calorie fast foods among the youth is compounding the situation. Most of these diseases are preventable if right intervention in terms of dietary pattern and regular physical activities are incorporated at the right age. ^35-40^

There have been several studies where the student perceptions about various aspects pertaining to their physical activities have been evaluated. Different approaches of inculcating the habit of leading a physically active lifestyle among the student community have also been suggested.^41-45^ However, the situation in India is complex owing to the presence of a variety of regulations and norms of developing curricula in higher education institutions. Similarly, there are different types of universities including deemed universities, private universities, state universities and central universities. The education policies so far have mostly emphasized on the importance of physical education in schools.^46^

Our study suggests that various curricula of higher education have several lapses when it comes to health-related topics. Universities need to take up the initiative in making the students aware of the correct ways of leading a healthy lifestyle. Irrespective of the stream of education, keeping oneself physically and psychologically fit is essential to lead a healthy life. Our results seem to suggest that health education must become a part of all streams of higher education irrespective of the stream.

## Conclusion

In our sample, we report that about 14.5% of all students fall under ‘Inactive’ category (14.4% among all males and 14.7% among all females), about 71.3% of all students (72.1% among all males and 70% among all females) fall under ‘Highly active’ category and about 14.2% of all students (13.5% of all males and 15.3% of female) fall under ‘active’ category. In our study we found that physical activity levels go on decreasing as the age increases, i.e., lowest physically active students belong to higher age group and highly active students are in lower age group. Our study also suggests that physical education and other aspects of health are inadequately and heterogeneously represented in university curricula. These topics are required to be incorporated into regular curricula in all streams of higher education in Indian universities.

## Data Availability

The original data reported in the manuscript is available with the corresponding author.

